# Multimorbidity in Older Adults with Depression Study (MODS) (Behavioural Activation to improve physical and mental functioning among older people with multiple long-term conditions): Protocol for a fully powered randomised controlled trial

**DOI:** 10.1101/2024.01.10.24301134

**Authors:** Eloise Ryde, Kelly Hollingsworth, Elizabeth Littlewood, Lucy Atha, Della Bailey, Heather Baker, Katharine Bosanquet, Leanne Shearsmith, Carolyn A. Chew-Graham, Dean McMillan, Kalpita Baird, Peter Coventry, Suzanne Crosland, Caroline Fairhurst, Han-I Wang, Lauren Burke, Janine Heeley, Catherine Hewitt, Andrew Clegg, Tom Gentry, Andrew Hill, Karina Lovell, Sarah Dexter-Smith, Gemma Traviss-Turner, Judith Webster, Elizabeth Agnew, Heidi Stevens, Katie Webb, Simon Gilbody, David Ekers

**Author notes:** Declared competing interests of authors: none.

## Abstract

**Introduction:** Older adults (65 years or over) and those with long-term health conditions (LTCs), represent a ‘high risk’ group for depression, with a risk two-to-three times the general population. This can lead to poorer quality of life and be costly to health and social care services. In the Multimorbidity in Older Adults with Depression Study (MODS) we will test whether a brief psychological intervention (Behavioural Activation), helps to improve physical/mental functioning in this group compared to treatment as usual.

**Methods:** We will conduct a two-arm, parallel-group randomised controlled trial, to evaluate the clinical and cost-effectiveness of the MODS intervention. Participants will be recruited via general practices across England. To be included, participants must be aged 65 years or over, with two or more LTCs and either sub-threshold depression or major depression. Randomisation will be simple 1:1. Intervention participants will receive up to eight sessions delivered by MODS support workers, supported by a self-help booklet. Control participants will receive usual care.

A process evaluation will be undertaken to evaluate the processes and mechanisms underpinning intervention delivery, and to inform the development of an implementation framework. Semi-structured interviews will be conducted with intervention participants, participant’s caregivers/supportive others, and health and social care professionals. Focus groups and semi-structured interviews will be conducted with MODS support workers. Outcome data will be collected at four, eight, and twelve-months post-randomisation. The primary outcome is self-reported quality of life and functioning at the four-month follow up. Secondary outcomes include depression, anxiety, physical functioning, loneliness, social isolation, chronic pain, health related quality of life, and health services use.

**Discussion:** This study builds on our previous work and will evaluate a brief psychological intervention to improve physical and mental health functioning for older adults with multiple long-term conditions.

**Trial Registration:** ClinicalTrials.Gov Identifier ISRCTN44184899, registered on 11^th^ August 2022.

## Introduction

With the UK population ageing, [1] and older adults representing a substantial and growing proportion of the global population, there is increased urgency to understand and address their unique physical health and mental health needs [2]. Older adults are a heterogenous population; it is not age alone that may create vulnerability, but the risk factors associated with ageing [3]. Along with the physical and social environments that may influence healthy ageing, personal characteristics, including the presence of long-term conditions (LTCs), may lead to a decline in physical and/or mental health functioning.

Having two or more LTCs is referred to by the National Institute for Health and Care Excellence (NICE) as ‘multimorbidity’ [4] though we will use the patient-preferred term ‘multiple long-term conditions’. Large-scale survey data (n=4,712) examining age-related change found that health-related variables (number of LTCs, self-rated health) were strongly associated with perceptions of physical functioning, which increased with age from 65 years onwards [5]. Yet the challenges of managing multiple LTCs are not exclusively physical.

Depression is two-to-three times more likely to be present across the range of LTCs resulting in poorer outcomes, lower quality of life and increased mortality [6]. Depression is defined as an emotional disorder characterised by the persistent experience of negative feelings such as sadness, emptiness, and joylessness, which is usually accompanied by lack of energy, tiredness, exhaustion, and fatigue [7]. Among older adults with co-morbid LTCs, depression is associated with the greatest decrements in quality of life, greater treatment costs and contributes to health inequalities, compared to those without LTCs [8]. In a healthcare context where the mental health needs of older adults are often unaddressed, and older adult psychological services are inadequately integrated across care settings [9], a growing body of research is exploring feasible and scalable psychological intervention development to address this unmet need.

There is accumulating evidence of cognitive and behavioural approaches in the prevention or mitigation of depression in older adults [10, 11]. Behavioural Activation (BA) is an evidence-based psychological treatment that explores how physical inactivity, avoidance, and low mood are linked, and result in a reduction of valued activity [12]. It aims to reinstate valued activities and connect individuals with sources of positive reinforcement. Moreover, there is evidence that BA is acceptable to older adults with LTCs [13]. Existing literature suggests that engaging in a greater variety of daily activities is related to increased social connectedness [14], which is protective against loneliness and mental ill-health. Additionally, there is evidence that a greater range and number of daily activities is related to higher psychological well-being among older adults, with increased diversity of activities over 10 years being linked to increased wellbeing [15]. Thus, brief psychological interventions aimed at increasing and facilitating older adults’ engagement in valued activities (such as BA) might have the potential to mediate the link between decline in physical functioning and depressive symptoms. A meta-analysis found that BA significantly reduced depressive symptoms in older adults in the community, but recommended further high-quality trials of BA for older adults with multiple LTCs are needed [11]. Whilst there is evidence demonstrating the clinical benefit of brief psychological interventions (including BA) for depression in the short term [16], the clinical and cost effectiveness of BA for older adults with multiple LTCs over the short and longer-term needs to be evaluated in a fully powered, randomised controlled trial (RCT), specifically evaluating both physical functioning and depression in older adults with LTCs.

The primary objective of the MODS trial is to conduct a fully powered, RCT to evaluate the clinical and cost effectiveness of a brief psychological intervention (BA), set within a collaborative care framework, for older adults with multiple LTCs and depression. A secondary aim is to inform post-trial implementation of the intervention across a range of provider settings and staff.

## Materials and Methods

### Research Aims

The aim of the MODS trial is to assess the clinical and cost effectiveness of a brief psychological intervention (BA delivered within a collaborative care framework), with embedded process, and economic evaluations:

- Establish the clinical and cost effectiveness of the MODS BA intervention compared to usual care on physical and mental functioning in older people with multiple LTCs.
- To conduct preliminary economic modelling of intervention effects.
- Evaluate the processes and mechanisms that underpin intervention delivery and to develop a post-trial implementation framework for use across provider settings and staff.

### Design

This is a two-arm parallel-group, multicentre, RCT with embedded qualitative process and economic evaluations. The trial also involves a sub-study to explore therapeutic alliance in brief psychological therapy. The process of participant enrolment, interventions and assessment is outlined in figure 1.

**Fig 1.**
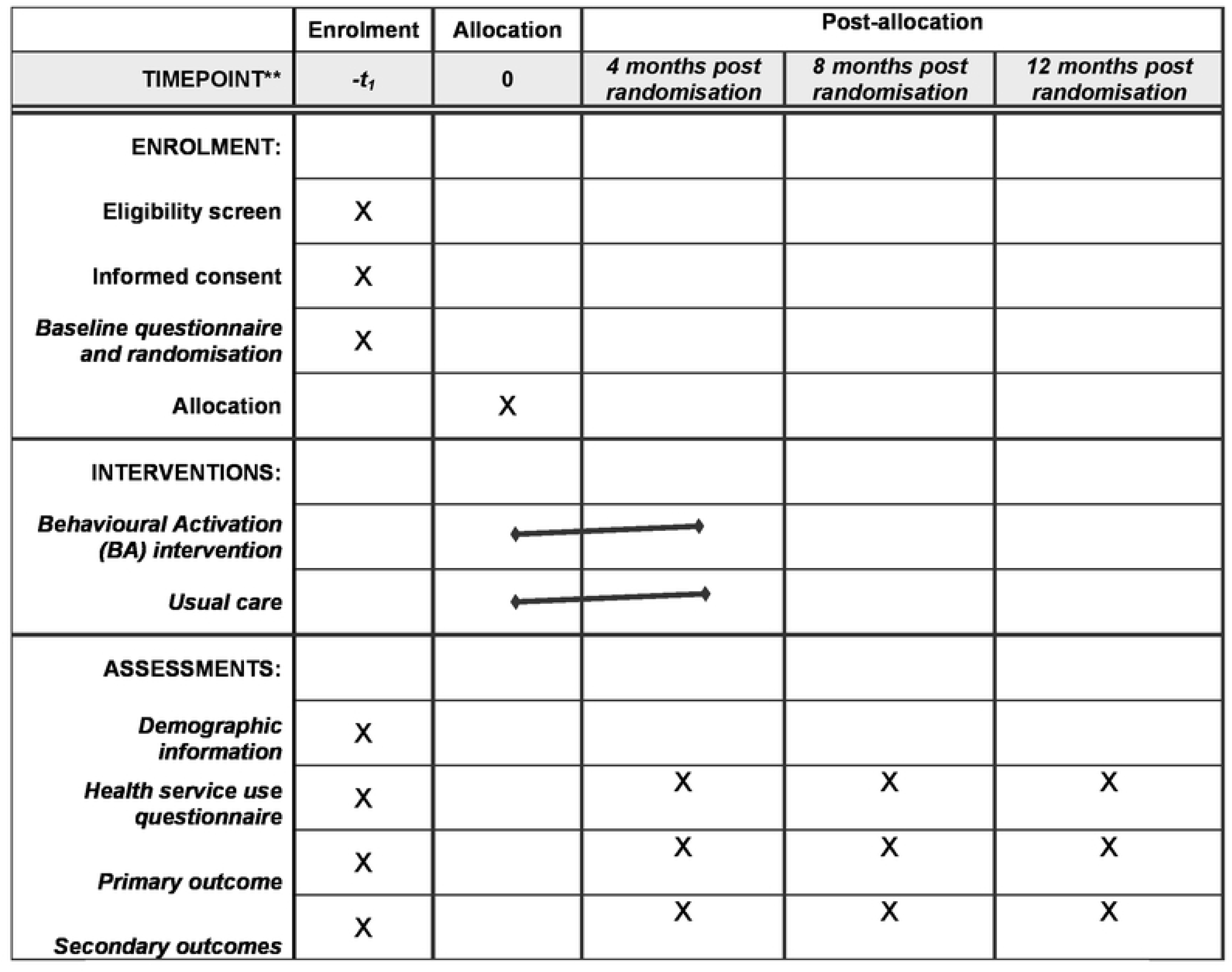
SPIRIT schedule of enrollment, interventions, and assessments.

### Setting

Participants will be identified through primary care general practices across England. The MODS BA intervention will be delivered across a range of health care settings (e.g., primary care, secondary care, and voluntary/third sector services).

### Identification

Potential participants will be identified through searches of general practice registers. Lists of patients aged 65 years or over with two or more LTCs will be screened by a member of the practice team to ensure patients meeting exclusion criteria (detailed below) are removed.

LTCs will be based on the Department of Health (DoH) definition [17] and will focus on commonly reported LTCs for older people (such as asthma/chronic obstructive pulmonary disease, diabetes, hypertension/coronary heart disease, and stroke) according to the primary care Quality and Outcomes Framework (QOF) [18] but will also include long-term conditions such as neurological conditions.

### Inclusion Criteria

- Older adults (65 years or over)
- Two or more long-term physical health conditions
- Sub-threshold or major depression as ascertained by the Structured Clinical Interview for DSM-5 axis 1 disorder depression subscale (SICD-5) [19]. Participants will be categorised with sub-threshold depression where 2-4 depression symptoms (where at least one of these are low mood or loss of interest or pleasure) are present. Where 5 or more depressive symptoms are present (where at least one of these are low mood or loss of interest or pleasure) participants will be categorised as having major depressive disorder.

### Exclusion Criteria

- Cognitive impairment
- Bipolar disorder/psychosis/psychotic symptoms
- Alcohol or drug dependence
- In the palliative phase of illness
- Have active suicidal ideation
- Currently receiving psychological therapy
- Unable to speak or understand English

Older adults will not be excluded based on living in residential/care homes.

### Recruitment

Potentially eligible participants will receive a study information pack (containing a practice letter-headed invitation letter, Participant Information Sheet (PIS), consent form, and a freepost return envelope) via their GP practice. Participants can indicate their interest in the study through completing and returning a written consent form using the freepost envelope, completing and submitting an online consent form, or by contacting the study team directly (study team contact details provided in the PIS). Where feasible, potential participants will be contacted by telephone, by extended GP practice teams, to establish interest in the study and gain ‘permission to contact’. Verbal permission to pass on contact details to the local MODS team will be recorded for interested patients.

Interested patients will then be contacted by telephone (or videocall, using an appropriate online platform) by a MODS researcher to discuss the study, answer questions, and assess eligibility. Verbal consent for study participation will be sought from interested and eligible participants where they have not fully completed a written/online consent form. This process (approved by the Research Ethics Committee) will involve the MODS researcher reading out each consent statement verbatim and asking the participant whether they agree or disagree with each statement, and documenting their response on a physical copy of the informed consent form. Once informed consent is confirmed/received, the baseline questionnaire will be completed over the telephone with a study researcher (either immediately following completion of the baseline questionnaire or at a later date, and preferably within one week of confirming eligibility, and in line with participant preference). Where participants may find completing the baseline questionnaire over the telephone difficult, the option to complete this via post (returned with a freepost envelope) or online (participants will be provided with a secure unique link to the questionnaire) will be considered..

### Randomisation and Blinding

Once the baseline questionnaire has been completed, eligible and consenting simple randomisation will be used to allocate participants 1:1 to either the BA intervention group or usual care group. Randomisation will be completed via a secure online randomisation service provided by York Trials Unit (YTU). A YTU statistician who is not involved in participant recruitment will generate the allocation schedule. Participants will be informed by telephone of their group allocation immediately after randomisation has taken place, and this will be confirmed by letter. For those participants allocated to the intervention group, a copy of the MODS intervention booklet will accompany their allocation letter. A letter will also be sent to the participant’s GP practice confirming inclusion in the study, group allocation and information regarding their mood (with the participant’s consent).

Researchers will be blind to a participant’s group allocation when completing follow up questionnaires (where these are completed over the telephone). Due to the nature of the intervention, it is not possible for those staff delivering the intervention (MODS Support Workers) or participants to remain blind to group allocation.

### The MODS Intervention

The MODS intervention utilises BA set within a Collaborative Care (CC) Framework. BA aims to maintain an individual’s connection with the world by helping them to continue with the activities they value. Where particular valued activities may no longer be possible, either temporarily or in the long-term, BA prompts participants to think about alternative activities which fulfil a similar function for them and help them to remain active. Remaining active and staying connected with the world may benefit physical and mental wellbeing. This may be particularly important as LTCs can restrict the activities a person is able to do and curtail their engagement with the outside world. The MODS Support Worker (MSW) and the participant work together using the MODS self-help booklet to develop an individualised treatment plan.

The CC aspect of the intervention involves the MSW encouraging and supporting the participant to take a proactive approach towards managing their mood and LTCs. The MSW will liaise with the participant’s GP or other professionals involved in their care, if appropriate, and where the participant consents to this. They may also signpost or help participants to access relevant support services or organisations, including those within the voluntary/third sector, who provide services or run activities which may be of interest to the participant.

Participants will be offered up to eight intervention sessions during which they will be supported by a trained MSW to work their way through the self-help booklet at a pace they feel comfortable with. The self-help booklet has been developed as part of the wider MODS research programme, with input and feedback gathered from a range of stakeholders, including older adults with LTCs and/or mental health conditions, and members of the MODS Patient and Public Involvement Advisory Group (PPI AG). For the majority of participants, sessions will be delivered remotely, either over the telephone or by video call, where this is available and according to participant preference. Face-to-face sessions may be offered on an individual basis where sessions over the telephone or via video call are not feasible; for example, where significant hearing difficulties make contact in this way difficult. The first session will last approximately one hour with subsequent sessions lasting approximately 30 minutes.

Symptom monitoring at each intervention session will be undertaken using the depression scale of the Depression Anxiety Stress Scale (DASS) [20]. The DASS is a widely used monitoring tool which is validated in a UK community context and is simple to score with clear and standard clinical cut off scores (non/mild/moderate/severe). DASS scores will be used to guide decision making by MSWs in conjunction with their MODS clinical supervisor. Where risk or significant clinical deterioration is indicated the MSW will support the participant to access more formal healthcare interventions.

### MODS Support Workers (MSWs)

MSWs will include a range of practitioners from across a variety of backgrounds, both clinical and non-clinical, and be based within primary care, secondary care, or voluntary/third sector settings. MSWs will be required to complete a remotely delivered bespoke intervention training course (approximately 22 hours) facilitated by clinical members of the MODS study team. Materials, including role-play demonstrations of sessions and a MSW treatment manual will be provided to the MSWs. The training will cover the components of the BA intervention set within the CC framework, intervention delivery including the MODS self-help booklet, and study procedures including those relating to managing risk and adverse events. MSWs will be required to pass a telephone-based bespoke competency assessment with a training facilitator before they commence delivery of the intervention. Regular supervision/support will be provided to the MSWs from a clinical member of the MODS study team.

### Comparator

Participants randomised to the usual care group will receive usual care as provided by their current NHS and/or third sector providers.

### Outcome Measures

Data will be obtained at baseline and four, eight, and twelve-months post-randomisation. Baseline data will be collected over the telephone with a researcher, while participants will have the option to complete follow-up questionnaires via the telephone (with a researcher), online (via a secure and unique link emailed to the participant), or via the post (with a free post return envelope provided). A reminder process consisting of emails and letters will be implemented, where appropriate.

The primary outcome will be self-reported quality of life and functioning measures (as measured by the mental and physical component scores of the SF-12v2) [21] at four months post randomisation.

Secondary outcomes will include quality of life and functioning (SF-12v2) at eight and 12 months; depression status according to DSM-5 criteria (SCID-5) [19]; depression severity (PHQ9) [22]; anxiety (GAD) [23]; physical function (NEADL) [24], loneliness (De Jong Gierveld Scale – 11 items, total score and the two subscales of Social and Emotional loneliness) [25]; social isolation (Lubben Social Network Scale - 6 items), chronic pain (two questions from the Graded chronic pain scale revised) [26], health related quality of life (EQ5D-3L) [27], and a bespoke health resource use questionnaire, each at four, eight, and twelve-months.

Demographic information, including age, LTC types/health condition(s), depression history, socio-economic status, ethnicity, education, cohabitation status, and Covid-19 history, will be obtained as part of the baseline questionnaire.

### Data Management Plans

All trial data will be securely stored on University or NHS computers. Remote access to data (for staff working remotely) will be via secure and approved organisational Virtual Private Networks (VPN), or equivalent. Where data is stored by non-NHS organisations, the process for data storage will be reviewed and approved by the trial Sponsor (Tees, Esk and Wear Valleys NHS Foundation Trust). All data storage processes will be in line with General Data Protection Regulation (GDPR) and Good Clinical Practice (GCP) guidance). Access to participant data will be restricted according to MODS researcher role. Participant confidentiality will be maintained throughout, unless significant risk to self or others is identified.

### Therapeutic Alliance Sub-Study

Research has shown the relationship between therapeutic alliance and therapy outcome in the treatment of depression is often a central component for the success of psychological therapies. However, much of the research supporting this claim examines the alliance-outcome relationship within high-intensity psychological treatments (such as Cognitive Behaviour Therapy), and currently little is known about therapeutic alliance within brief psychological treatments such as BA.

A sub-study examining therapeutic alliance will be incorporated within the MODS study to allow exploration of if, and how, therapeutic alliance may predict intervention outcomes (such as depression score) in brief psychological treatments.

A measure of therapeutic alliance (the Agnew Relationship Measure 12 item (ARM-12), [28]) which gathers information on the strength of therapeutic alliance between the MODS support worker and participant, will be incorporated into the MODS intervention. Participants randomised to the MODS BA intervention group will receive blank printed copies of the ARM-12 measure (the participant version) alongside their allocation letter, associated intervention materials, and a freepost return envelope. The allocation letter details how and when to complete the ARM-12 measure. MODS Support Workers will have the option to complete their version of the ARM-12 measure on hard copies or online via a secure link.

Both the participant and the MODS Support Worker will complete the ARM-12 measure independently following each MODS BA session. A latent trajectory analysis will be conducted where sufficient ARM-12 data is collected, as this will allow for change over time assessment. If this analysis is not feasible, a multi-level regression may be conducted. This therapeutic alliance sub-study will be reported separately to the statistical analysis detailed later in this paper.

### Safety Considerations

Participant risk (suicide and non-suicide) will be monitored by study researchers and MSWs during all participant contacts. Standard operating procedures and risk assessment training will be provided. Where risk is identified, clinical members of the MODS study team will support the risk assessment and determine the level of risk and, where appropriate, will provide information to GP practices or emergency services.

Serious adverse events and adverse events will also be monitored by study researchers and MSWs. These events will be reported within the appropriate timeframe.

### Sample Size

To detect a small to medium standardised effect size of 0.3, on either the mental or physical component score of the SF-12v2, assuming an alpha level of 0.025 and 80% power, a total sample size of 426 participants is required. An effect size of 0.3 corresponds to a difference of 3.3 SF-12v2 score points, assuming a standard deviation of 11 [29, 30], which falls within the range of estimated minimum clinically important differences for SF-12v2 from varying populations [31]. Although this is an individually randomised trial, the sample size was inflated to account for potential clustering effects within MSWs, based on an intracluster correlation coefficient (ICC) of 0.01 (in line with empirical estimates of within-therapist clustering obtained in the CASPER trial [30] and an average cluster size of 15 (design effect = 1.14). Though there is only clustering by MSW in the BA intervention group, the adjustment was made for both groups, which provides a more conservative sample size target. Allowing for 15% attrition, 572 participants, approximately 286 in each arm, would need to be recruited and randomised into the trial.

### Analyses

#### Statistical Analyses

A detailed statistical analysis plan (SAP) will be produced before data analysis commences. This will be approved by the joint Programme Steering and Data Monitoring and Ethics Committee. Analysis will be conducted on an intention to treat (ITT) basis, using two-sided statistical tests at the 5% significance level, using Stata v17 or later. The statistician will not be blinded to treatment allocation.

The flow of participants through the trial will be presented using a CONSORT diagram [Figure 2]. This will include the number of individuals screened, eligible and randomised with reasons for non-participation provided where available. Adherence to the intervention will also be recorded and reported. Full withdrawal and intervention only withdrawal will be summarised according to trial arm.

**Figure 2:**
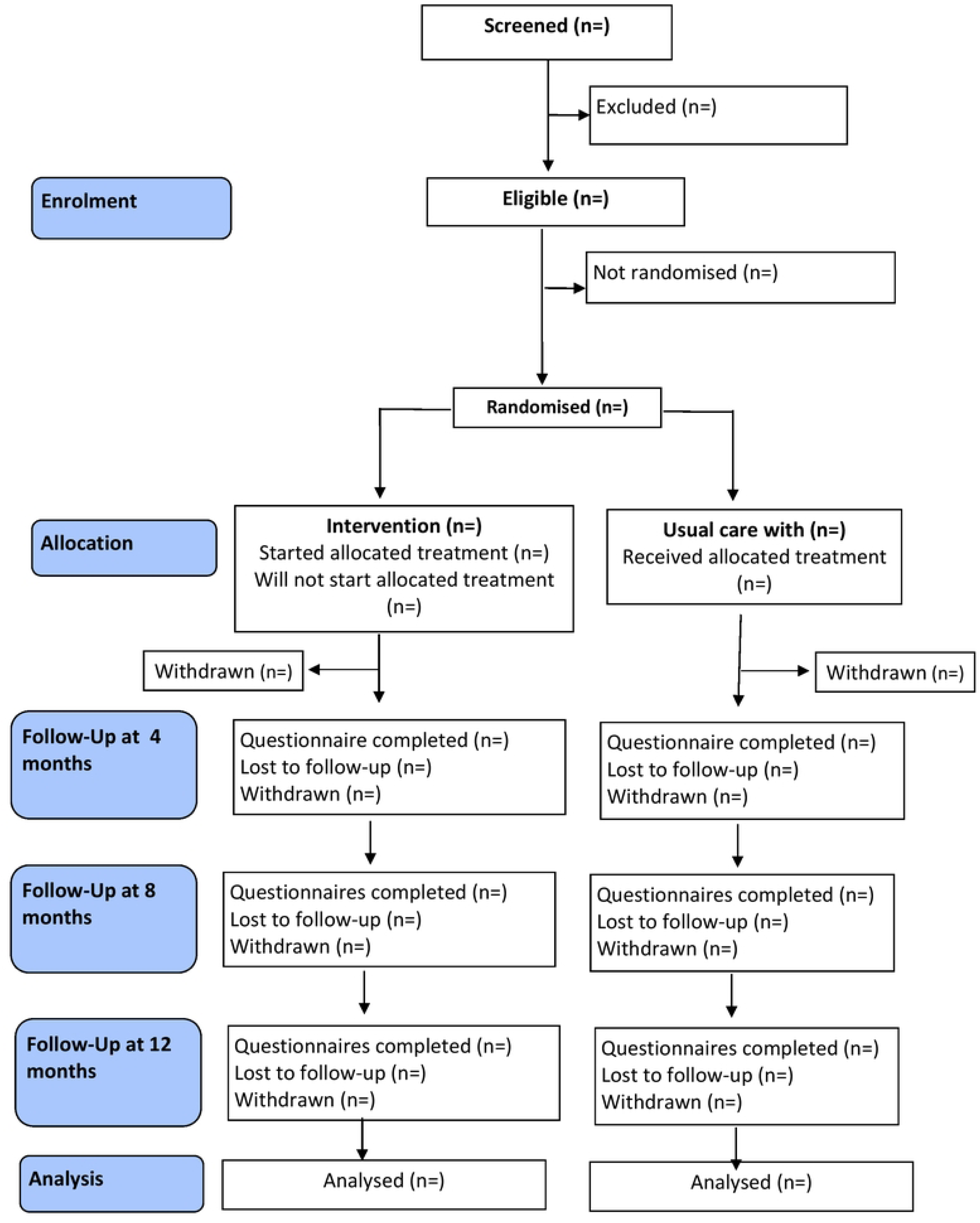
MODS CONSORT flow diagram.

Baseline data will be summarised by trial arm for all participants as randomised and as included in the primary analysis. Formal statistical comparisons will not be completed on baseline data. Continuous measures will be reported as means and standard deviation (SD), while categorical data will be reported as counts and percentages.

The primary outcomes (Physical and Mental Component Scores of the SF-12v2) will be analysed separately using a linear mixed model, including assessments at all available follow-up time points (four, eight and twelve months after randomisation). The model will adjust for baseline value of the outcome measure, trial arm, time, and arm by time interaction as fixed effects. Random effects will be participant, MSW, and site. The model will provide an overall treatment effect over 12 months, as well as estimates at individual time points, which will be reported as adjusted mean differences with associated 95% confidence interval (CI) and p-value. The primary time point of interest is four months.

A complier average causal effect (CACE) analysis will be conducted for the primary outcome to account for non-compliance with the intervention. Exploratory subgroup analyses for a range of moderators of effect for the primary outcomes at four months (e.g. LTC type, age, depression history, socioeconomic status) will be undertaken.

Secondary continuous outcomes (PHQ9, GAD7, NEADL, De Jong Gierveld Scale [Social loneliness subscale, Emotional loneliness subscale and overall], Lubben Social Network Scale, Graded chronic pain scale revised) will be analysed using the same methods as described for the primary analyses. The categorical outcome of depression status as identified by the SCID-5 will be analysed using logistic regression and presented using odds ratios, 95% CIs and p-values.

#### Economic Analyses

The primary analysis of the economic evaluation will evaluate the cost effectiveness of the MODS BA intervention compared to usual care for older adults with multiple LTCs and depression from a National Health Service (NHS) and Personal Social Services (PSS) perspective. The cost of the intervention will be obtained via the MODS team; while costs of health and social service use data will be obtained from participants via a self-completed, brief, bespoke questionnaire administered at each follow up. Health outcomes will be measured in terms of quality-adjusted life years (QALYs) using the EQ-5D-3L questionnaire and calculated using standard area-under-the-curve method. The differences in costs and QALYs between the intervention and usual care groups, adjusted for baseline characteristics, will be used to calculate the incremental cost-effectiveness ratio (ICER) against the willingness-to-pay threshold in the UK. Uncertainties around the estimated ICER will be explored using non-parametric bootstrapping methods with 5,000 iterations. The results will be presented graphically on the cost-effectiveness plane and cost-effectiveness acceptability curve. Sensitivity analyses will be conducted to test the robustness of the cost-effectiveness results under various scenarios.

To assess the long-term cost effectiveness of the intervention beyond-trial evaluation using model-based approach will be explored and considered if the within-trial evaluation results deem appropriate (e.g. the intervention is not dominant by usual care). For the projection, a decision model will be created using evidence from the MODS trial and the wider published literature to produce an estimate of the long-term health outcomes and health care costs. Probabilistic sensitivity analyses will be conducted to assess the robustness of the model results.

### Ethical Considerations and Declarations

The MODS trial received ethical approval from the Yorkshire and The Humber – Leeds West Research Ethics Committee on 27^th^ May 2022 (REC Ref: 22/YH/0071). The sponsor for MODS is Tees, Esk and Wear Valleys NHS Foundation Trust.

Although our study population could be considered to be vulnerable, we do not foresee any major ethical issues. Protection of the human rights and dignity of participants will be in place during the trial, in line with the 1996 Helsinki Declaration. The study has been designed to minimise any risk for the participants when taking part in the study. Participants’ wishes will be respected at all times, including the right to withdraw from the study at any time without giving a reason. The interests of the patient will be held above those of science and society and provision will be made for indemnity by the investigator and sponsor. Care that is currently available via the NHS will not be withheld from participants.

Protocol amendments will be managed via the Health Research Authority, Research Ethics Committee, and sponsor approvals process throughout the duration of the study.

### Qualitative Process Evaluation

The qualitative process evaluation will explore the impact of the intervention on the physical and mental functioning of MODS intervention participants. It will also explore pathways to implementation by seeking to identify possible barriers and enablers to the delivery of the intervention in practice, beyond the confines of a research study.

Approximately 20-25 semi-structured interviews will be conducted with intervention participants, to include those who declined the intervention or who started sessions but disengaged (‘non-completers’); and those who completed the intervention (‘completers’). Consent to take part in an interview will be obtained as part of a set of optional consent statements upon study entry.

We will also conduct approximately 10 semi-structured interviews with caregivers or the supportive others of intervention participants. Participants and MSWs will identify potential caregivers/supportive others and consent will be sought to provide the MODS team with contact details for sending a caregivers/supportive other information pack (containing a study invitation letter, PIS, consent form and freepost envelope). Interested caregivers/supportive others will complete a written consent form or verbal consent will be taken prior to conducting the interview.

Interviews with participants and caregivers/supportive others will be conducted over the telephone or via a virtual platform and last up to approximately 45 minutes. All interviews will be conducted after completion of the primary outcome.

MODS support workers will be purposively sampled to include a range of characteristics, including service/organisation type, job role, site and years of service. Three or four online focus groups will be held with approximately 20 MSWs. Each group will include four to six MSWs, ideally from different recruiting sites. MSWs will also be given the option of taking part in one-to-one telephone interviews where they are unable to join a focus group, or if this is their preferred method of providing feedback. MSWs will be invited to indicate their interest by contacting the study team, discussing the opportunity with their MODS clinical supervisor or by completing an online consent form. The focus groups will last approximately 45-60 minutes and will be conducted via an online platform. Semi-structured interviews will be conducted by telephone or video call and will last around 30-45 minutes.

We will also aim to interview a cross-section of approximately 10 health and social care professionals (e.g. GPs, hospital practitioners, social care managers). MSWs will help identify these professionals where they have had contact as part of the collaborative care aspect of the intervention. Interested health and social care professionals will be sent a study information pack (containing a study invite letter, a PIS and a consent form) and invited to complete an online consent form to register their interest.

Interviews/focus groups with the four participant groups (intervention participants; caregivers/supportive others; MSWs; health and social care professionals) will be conducted in parallel so that data analysis in each dataset enables modification of topic guides as the study progresses, as appropriate. Interview topic guides will be tailored to each participant group. Final numbers of participants will be determined by achievement of data saturation in each dataset [32].

All interviews/focus groups will be digitally recorded (with participant consent), anonymised and transcribed using a professional transcription service, with the transcripts forming the raw data for analysis. Initially, thematic analysis [33] will be conducted using a framework approach [34]. A coding framework will be developed, where codes will be examined across individual transcripts as well as across the entire data set and allocated to the framework.

Using aspects of the constant comparison method of analysis [35, 36], broader categories using linking codes will be developed across the transcripts. Further analysis will be guided by Normalisation Process Theory (NPT) [37] framework to structure participants’, caregivers/supportive others’ and health and social care professionals’ views about acceptability and implementation of the intervention and how it might be implemented in routine services.

### Patient and Public Involvement

The MODS PPI AG was convened in 2018 at the start of the MODS programme of research. The group currently consists of seven individuals with a range of lived experience (including older adults with physical-mental comorbidities) and caregivers/supportive others, and includes the MODS PPI Co-Investigator.

The role of the PPI AG is to support the MODS research programme. Members of the research team and the PPI AG have met on numerous occasions both in person and virtually to discuss study procedures and materials. The PPI AG provided feedback on many aspects of the design and delivery of the MODS RCT; this included the use of postal consent forms and the development of recruitment, intervention, and participant materials. Importantly, they provided guidance and advice on how best to engage older adults with LTCs and their caregivers (where identified). The PPI AG will also be involved with the dissemination strategy, to ensure the findings are accessible to a range of audiences, including study participants and the public.

Our PPI Co-Investigator (JW) is part of our wider research team, and attends Programme Management Group meetings, contributes to ongoing discussions relating to the progression of the research programme and liaises with our Age UK partner. Two members of the PPI AG also attend Programme Management Group meetings; this is done on a one-year term so that each member of the PPI AG has the opportunity to attend Programme Management Group meetings should they wish. This format was discussed with and agreed by the PPI AG members. In addition, an independent PPI representative sits on the joint Programme Steering Committee and Data Monitoring & Ethics Committee to provide PPI input for the entirety of the programme.

### Study Status and Timeline

Recruitment opened on 13^th^ July 2022 and was estimated to be completed by 29^th^ February 2024. Due to ongoing recruitment challenges, and following an unsuccessful application to the funder to extend the study to meet the required sample size, recruitment will now cease earlier than planned, closing on 21^st^ December 2023. Intervention delivery is expected to finish by April 2024. Follow up data collection will end in May 2024. The process evaluation will be completed in full. The current MODS protocol is version 3.0 dated 8th June 2023.

## Discussion

Older adults with two or more LTCs are at an increased risk of developing depression. NICE highlights the need for research to evaluate care packages that are tailored to an individual’s physical and/or mental health needs, and which optimises services for older people with multiple LTCs. MODS has been designed to respond to this growing need to address the impact of multiple LTCs in older adults by targeting both physical and psychological conditions within the same intervention.

BA is an evidence-based brief intervention that has been shown to reduce symptoms of depression in older adults [11]. We adapted BA for use with older adults with both physical and mental health conditions as part of the wider MODS programme. The MODS intervention has been designed to be delivered by staff from a range of backgrounds and to be delivered remotely.

The study has the potential to generate an effective care package which can be scaled up for delivery across a range of settings, leading to significant benefit to the NHS and community-based settings. Despite this, recruitment to the study proved challenging, mostly likely related in part to the current and ongoing pressures within the NHS, particularly within primary care. To this end, the decision was taken (by the funder following an unsuccessful application to extend the study duration) to cease recruitment short of our estimated required sample size.

The detailed process evaluation will be completed in full and will provide rich data on the impact of the intervention on the physical and mental health of older adults; important data will also be generated to inform pathways to implementation of brief interventions to support future research in this area. The delivery of the MODS RCT and its associated recruitment challenges has provided important learning opportunities which will inform future mental health research, especially where this involves recruitment of participants via NHS settings.

## Data Availability

No datasets were generated or analysed during the current study. All relevant data from this study will be made available upon study completion.

## Contributions of Authors

**Conceptualization:** David Ekers, Simon Gilbody, Dean McMillan, Carolyn A. Chew Graham, Peter Coventry, Catherine Hewitt, Gemma Traviss-Turner, Karina Lovell, Sarah Dexter Smith, Andrew Clegg, Tom Gentry, Andrew J Hill, Elizabeth Littlewood, Della Bailey, Judith Webster

**Data curation:** Caroline Fairhurst, Kalpita Baird, Catherine Hewitt, Katharine Bosanquet, Elizabeth Littlewood, Lucy Atha, Lauren Burke, Leanne Shearsmith, Han-I Wang

**Formal analysis:** Caroline Fairhurst, Catherine Hewitt, Kalpita Baird, Carolyn A Chew-Graham, Peter Coventry, Katharine Bosanquet, Han-I Wang

**Funding acquisition:** Simon Gilbody, David Ekers, Carolyn A. Chew-Graham, Dean McMillan, Catherine Hewitt, Peter Coventry, Gemma Traviss-Turner, Karina Lovell, Sarah Dexter Smith, Andrew Clegg, Tom Gentry, Andrew J Hill

**Investigation:** Eloise Ryde, Kelly Hollingsworth, Elizabeth Littlewood, Della Bailey, Lucy Atha, Leanne Shearsmith, Katharine Bosanquet, Dean McMillan, Lauren Burke, Suzanne Crosland, Heather Baker, Janine Heeley, Elizabeth Agnew, Heidi Stevens, Katie Webb

**Methodology:** Elizabeth Littlewood, David Ekers, Simon Gilbody, Carolyn A. Chew-Graham, Dean McMillan, Peter Coventry, Caroline Fairhurst, Catherine Hewitt, Katharine Bosanquet, Han-I Wang, Gemma Traviss-Turner, Andrew Clegg, Andrew J Hill, Karina Lovell

**Project administration:** David Ekers, Simon Gilbody, Elizabeth Littlewood, Lucy Atha, Dean McMillan, Lauren Burke, Della Bailey, Carolyn A Chew-Graham, Peter Coventry, Katharine Bosanquet, Han-I Wang

**Resources:** Dean McMillan, Della Bailey, Carolyn A Chew-Graham, David Ekers, Caroline Fairhurst, Kalpita Baird, Catherine Hewitt, Han-I Wang

**Software:** Caroline Fairhurst, Catherine Hewitt, Kalpita Baird, Han-I Wang

**Supervision:** Simon Gilbody, David Ekers, Dean McMillan, Carolyn A. Chew-Graham, Peter Coventry, Elizabeth Littlewood, Della Bailey, Suzanne Crosland, Lucy Atha, Lauren Burke

**Validation:** David Ekers, Simon Gilbody, Caroline Fairhurst, Kalpita Baird, Catherine Hewitt, Carolyn A Chew-Graham, Peter Coventry, Katharine Bosanquet, Han-I Wang

**Visualization:** Eloise Ryde, Kelly Hollingsworth, Elizabeth Littlewood

**Writing – original draft:** Eloise Ryde, Kelly Hollingsworth, Elizabeth Littlewood, Leanne Shearsmith, Della Bailey, Heather Baker, Lucy Atha, Katharine Bosanquet, Caroline Fairhurst, Kalpita Baird, Han-I Wang

**Writing – review & editing:** Eloise Ryde, Kelly Hollingsworth, Elizabeth Littlewood, Della Bailey, Heather Baker, Lucy Atha, Katharine Bosanquet, Leanne Shearsmith, Caroline Fairhurst, Han-I Wang, David Ekers, Simon Gilbody, Carolyn A. Chew-Graham, Dean McMillan, Peter Coventry, Andrew J Hill, Andrew Clegg, Suzanne Crosland, Judith Webster, Catherine Hewitt, Gemma Traviss-Turner, Karina Lovell, Sarah Dexter Smith, Tom Gentry, Lauren Burke, Katie Webb, Heidi Stevens.

## Acknowledgements

We would like to thank: (1) all the participants for taking part in the trial, (2) general practice and Local Clinical Research Network staff for identifying and facilitating recruitment of participants, (3) research sites and intervention delivery staff for aiding with study delivery and follow up activities, (4) the independent joint Programme Steering and Data Monitoring and Ethics Committee members for overseeing the study, and (5) our PPI AG members for their continued support, guidance and collaboration.

## Supporting Information

S1 File. SPIRIT checklist. (DOC)

S2 File. MODS WS3-4 Trial Protocol v3.0 08.06.23. (PDF)

